# The contribution of social and family networks in supporting care of children with sickle cell disease in Tanzania

**DOI:** 10.1101/2022.11.01.22281740

**Authors:** Daima Bukini, Irene Msirikale, Collins Kanza, Jesca Odengo, Deogratius Maingu, Upendo Masamu, Karim Manji, Julie Makani, Columba Mbekenga

## Abstract

**Background:** The care of children with Sickle cell disease (SCD) in Africa has been the work of mothers or women within communities. Previous studies have indicated that mothers in these families are supported by other women within the family or even from outside family structures. Such support has allowed significant sharing of responsibilities in both domestic and care giving roles for children with sickle cell disease. However, there is limited recognition of this informal support and its importance as a key element in health care provision.

**Objective:** In this paper, we aim to demonstrate how these support networks have been used by mothers in shouldering their caregiving roles. We also propose how the networks can be integrated into the health care system to improve quality of care of the children with SCD.

**Methods:** We conducted interviews with families of children who are diagnosed with sickle cell disease through newborn screening program in Tanzania between 2015 and 2019. In total, 15 families were included through focus group discussions (FGDs), dyadic interviews, and individual interviews. Also, a survey was conducted involving 100 families of children with SCD aged between 0 and 17 years to assess the different networks of care that exists within families and communities. Thematic analysis was used for the qualitative data while data from the survey was presented in proportions as pie charts.

**Results:** The results from the FGDs and IDIs demonstrated three ways in which the networks of care were supportive to mothers (1) Facilitating sickle cell disease diagnosis to children who were undiagnosed (2) Caring for the sick child (children) after diagnosis and (3) Support at home in caring for other children who are not sick and helping with domestic work responsibilities. Survey results indicated that the highest proportion of the respondents listed mothers of the children as the primary care giver (88%), followed by grandmothers (10%) and close family members (2%). Extended family members (20%) were recognized as the largest network of care in the absence of the primary caregiver outside the parents, followed by siblings, defined as elder brothers and sisters (18%) and grandparents (16%).

**Conclusion:** The findings from this study confirm that there is an important network supporting care of children with SCD in communities supporting parents. Enabling these support networks to be more formally integrated into the health care system will ensure those members of the community providing care are equipped with knowledge on sickle cell disease and having positive impact on the quality of care of the children born with SCD in sub-Saharan Africa.

## Introduction

Some of the proposed approaches to reduce stigma and burden of care on mothers with SCD children in Africa is to design strategies aiming to involve men in the care of these children (Bukini et al., 2020; Gervais et al., 2015). While this proposal is central as a step to reduce the gendered burden of care, we also believe through interrogating culturally relevant gender roles and responsibilities in the settings will also help to inform a successful public health intervention framed in a culturally appropriate manner (Berghs et al., 2020; Brown, 2012; de Vries et al., 2020; Dennis-Antwi et al., 2011; Marsh et al., 2011). Gender roles in different societies are uniquely informed by social norms and values of the society itself (Cornwall, 2005; Doucet, 2015). The definition of “family” in western contexts typically includes a mother, father, and children (i.e., *nuclear family*. In African settings, a family typically includes extended family members and, largely, even the community (Amoateng et al., 2007). Caregiving for sick children in African contexts is regarded as not only the responsibility of the mother or and the father, but also extended family members and the community at large. The informal SCD support networks discussed here have been key in shouldering the burden of care for mothers of children with SCD. Afterall, Evidence on the impact of social capital in health promotion is well recognized. (Barbarin et al., 1999; Mbekenga et al., 2011; Nieminen et al., 2013; Noll et al., 1994).

Borrowing from the scholars of African indigenous ethics, often referred to as Ubuntu, this may not come as surprise because it is already known that families in these contexts are communal in nature (Behrens, 2013; Marshall & Koedg, 2004). This practical connection of theories and practices show the potential of using locally relevant theories and practices in trying to address contextual issues in clinical practices in Africa. SCD, as the most common genetic disease in Africa, is a model disease that will significantly benefit if care interventions shift attention to understand what is happening at the community level and who is involved in and outside the hospital. Using the case studies from the newborn screening for SCD cohort, we attempt to show how the existing gendered networks of care in our communities can be further developed and adapted to create locally appropriate genetic intervention programs.

## Methods

### QUALITATIVE DATA: *FOCUS GROUP DISCUSSIONS (FGDs) AND IN-DEPTH INTERVIEWS (IDIs)*

Data used were part of a qualitative study that aimed to show the impact of the disproportionate burden of care to mothers of children with SCD and the effects on the quality of care (Bukini et al 2020). The study used purposive sampling through FGDs and IDIs, to develop an in-depth understanding of gender norms in this setting and how it influences childcare. For this article, we analyzed data related to what networks of care exists within our communities in supporting care of children with SCD in families, specifically how they have helped reduce the burden of care to mothers. The interviews and FGDs discussions were with families of children who were diagnosed with SCD through newborn screening (NBS) programs in Tanzania between 2015 to March 2019. In total 15-families were included as indicated in Figure 1 below. The analysis started from the codebook of the previous analysis using thematic content analysis. We have presented the data from the interviews as case series narrating ways on how networks of care within the communities’ support care of children with SCD.

**Figure 1.**
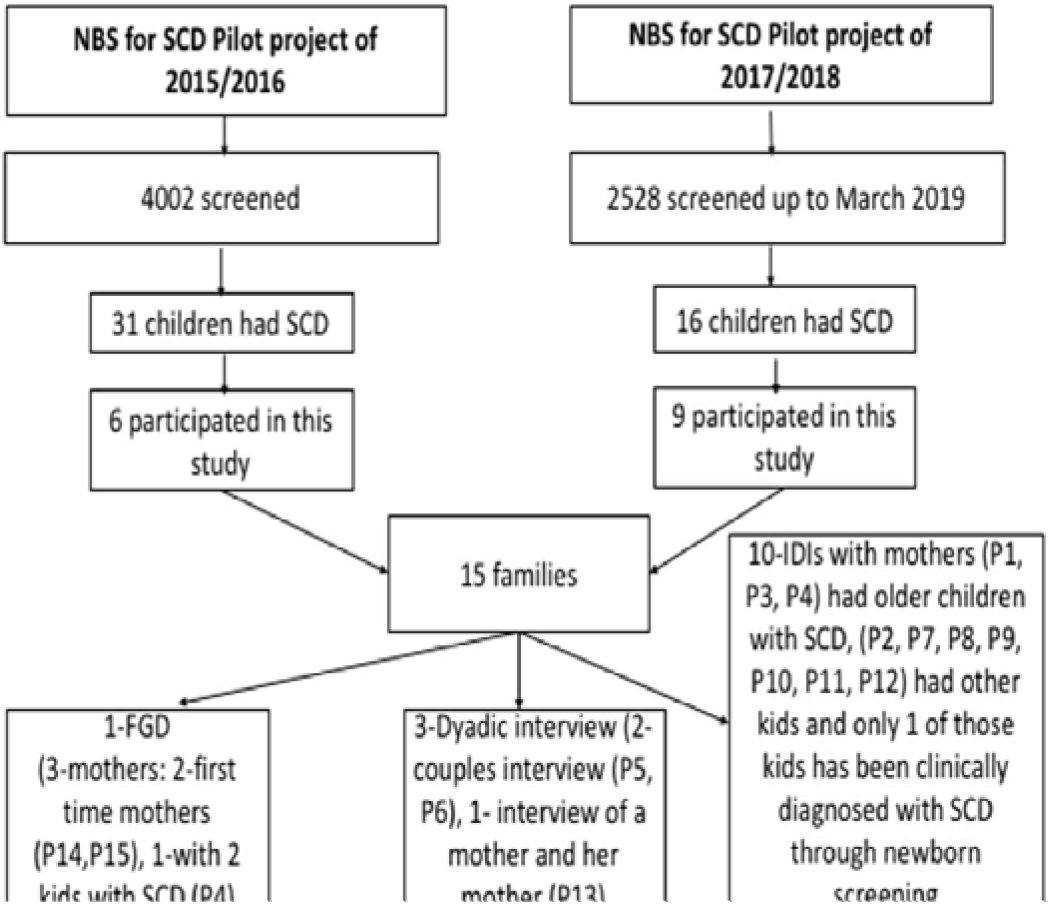
Recruitment flow chart for the 15-families who participated in the study (Bukini et al, 2020)

### QUANTITATIVE DATA: *SURVEY*

We developed a survey to further provide insights of the findings from the qualitative study with a larger sample size. In total 100 families of children aged 0-17 years of age attending SCD clinics in Dar es Salaam between November 2021 and February 2022 participated in the survey. The survey questions were developed from the themes identified from the qualitative data. Some of the questions in the survey included [a] who has been considered as a primary caregiver of the child? who is providing support when sick or hospitalized? [b] In the absence of the primary care giver who is providing support. Who has the responsibility of accompanying the child to the clinics for normal routine checkups or when sick?

## Results

### Qualitative

We inventory below ways in which extended family members and networks have supported mothers with sickle cell disease children.

#### 1) Support in facilitating SCD diagnosis within the communities

During the interviews, we observed how mothers with SCD children have been acting as facilitators for new SCD diagnosis within the community. Some cases of SCD diagnosis initiation starts at community level supported through women who either have children with SCD or had experience with caring for SCD patients. These women include family members, neighbors, or friends. The support starts by encouraging mothers to take their child/ children to hospitals for SCD testing and in some cases offering to pay for the expenses related to the diagnosis (transport, consultation, or diagnostic costs). *Case 1* below exemplifies how a mother came to diagnose her children with SCD and support that she did receive throughout her journey from diagnosis and care. Her previous experience with SCD was also instrumental in initiating newborn screening for her child and for other children who were delivered on that day.

##### Case 1

“*Mother of twins, both diagnosed with SCD after they started developing complications related to SCD. Her journey that leads to diagnosis was through a neighbour who had a child with SCD. She offered to take the mother to a local private hospital where the twins were diagnosed with SCD. In her next pregnancy, the mother requested for her child to be screened for SCD, This request did prompt the nurse to screen other mothers who also agreed for their children to be tested for SCD”*.

#### 2) Supporting care of the child or children after diagnosis

The care here includes care support after the children have been diagnosed with SCD. In most of the cases, mothers of children with SCD were either supported by women in their families to take children to the clinics for follow-up visitations, or support when children and mothers are hospitalized. Other forms of support were through help with the domestic work when the mother is at the hospital either for routine check-ups or hospitalization. It is worth noting that apart from the burden of caring for a child or children with SCD, most of these mothers have other responsibilities at home. These may include taking care of other children who may not have SCD but also domestic responsibilities at home such as cooking, cleaning, and washing.

##### Case 2

*“Participant was a mother who had recently had her newborn baby diagnosed with SCD through a local hospital. The mother has 4 children in total, with family history of SCD. The mother has responsibilities of taking care of all the four children. The mother explained that she is mostly relying on relatives to assist with bringing food at the hospital when hospitalized and other relatives assisting at home with the domestic work. She also acknowledged support she received from her husband to support the health care costs for the children with SCD”*.

#### 3) When assisted in caregiving, mothers can engage in economic activities to support children’s health care costs

We have also observed that for mothers who had support at home in caring for the children and in doing the housework, they managed to engage in economic activity outside their homes to help support themselves and their children. The support provided to the mothers enabled them to cover health care related costs, such as transport costs to and from the hospitals, paying for health insurance, or covering medical expenses at the hospital.

##### Case 3

*“Participant was a mother of three children, the youngest with SCD. Family with a history of SCD, husband did come for the genetic counselling session/ health education after the results which indicate support. The mother is currently doing small business at home, and while working, her other kids are being looked after by relatives. Without the support she is receiving from her family in caring for her children, she will not be able to engage in small business at home”*

### Quantitative

Results from the survey referenced above were categorized into four groups.

#### (1) Who is considered as primary caregiver of the child?

The highest proportion of the respondents listed mothers of the children as the primary care giver (88%), followed by grandparents (10%) and close family members (2%). These results complemented the findings from the qualitative work that there is disproportionate burden of care to the mothers, and that support is coming from mostly women family members, in this case, the grandmothers.

#### (2) Who has the responsibility of accompanying the child to the SCD clinics for normal routine checkups or when sick?

Sixty one percent (61%) of the respondents listed parents, defined as either mother or father or both; 12% listed extended family members, defined as aunts and uncles; 9% of the respondents listed elder sisters and brothers; 9% of the respondents listed grandparents; 8% came alone, these were some of the children aged 14 years and above; 1% listed other community members such as neighbors.

#### (3) In the absence of the primary caregiver who is providing support to the child at home or when sick or hospitalized?

**R**esponses were as follows: 45% listed parents, followed by extended family members (20%), siblings defined as elder brothers and sisters (18%), grandparents (16%), Community members defined as those with no family links such as neighbors (1%). Level of non-respondents for this question was 1%.

#### (4) Who is shouldering the domestic responsibilities when the primary caregiver is caring for the sick child or children in the hospital during clinics or when hospitalized?

Responses were as follows: 28% listed parents; 28% listed extended family members; 23% of the respondents listed grandparents, 18% listed siblings, for 3% of the respondents this question was not relevant and 1% listed other members of the community.

Figure 2 below are pie charts showing percentage distribution of the responses across the four identified categories.

**Figure 2.**
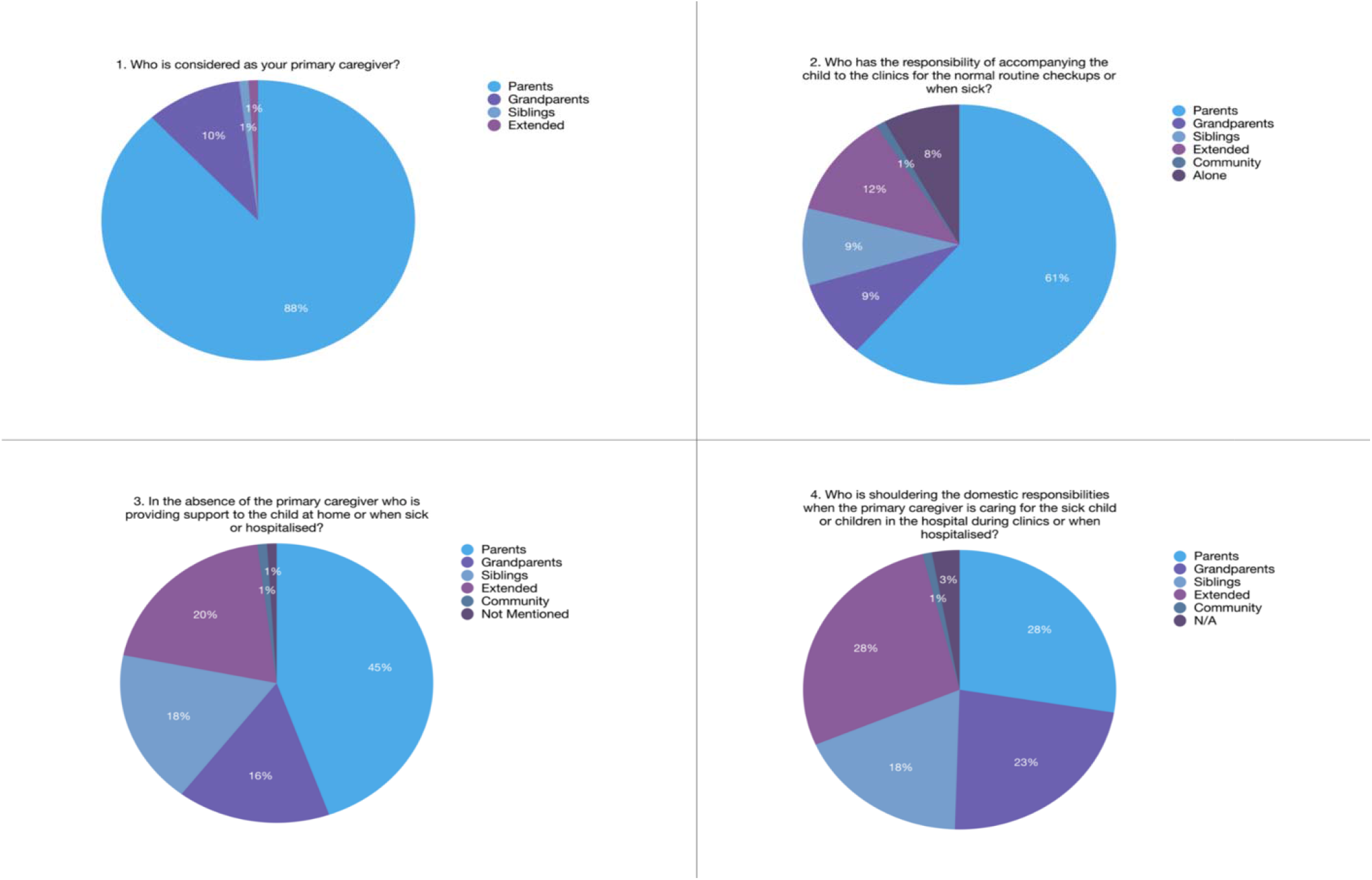
pie charts showing percentage distribution of the respondents based on the four main categories of questions as referenced in the results section

In summary, the findings from the survey as presented by the four categories of have shown that the networks of care within the community expand from the family unit (consisting of parents) to close family members (grandparents and other relatives) outward to include extended family members to those with no family links such as neighbors. The observation was consistent with the findings from the qualitative analysis that mothers of children with SCD receive support in caring for the children from different sources within families and outside the families. Figure 3 exemplifies the expansion of care from Part A to Part D.

**Figure 3.**
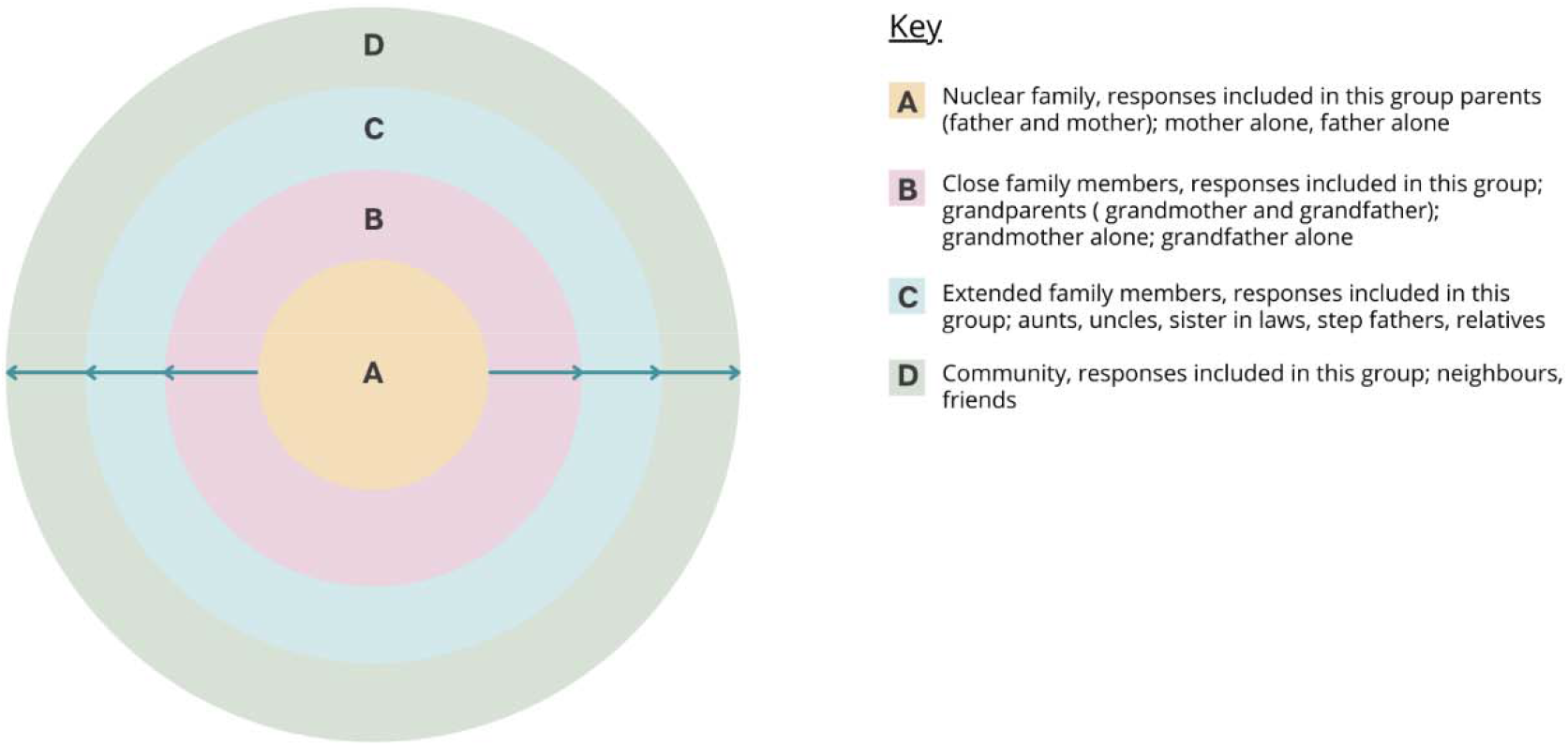
presenting the aggregate summary of the different networks of care existing within the families and outside in supporting care for children with SCD

## Discussion

This study used mixed methods design to provide evidence on how care for children with SCD extend from being the responsibility of the mother and in some instances the father to being a shared responsibility within communities. Other studies have suggested how the care burden can have negative impact on the families, especially mothers, hence affect quality of care provided to the child (Craft-Rosenberg et al., 2012; Olwit et al., 2018; Van Den Tweel et al., 2008). The support received from different parts of the networks of care, as shown in this study, provide significant contribution in supporting care of the children in and outside the clinics. If the structure and mechanisms of how these networks of care are well studied and integrated within the health care system, there is potential to build a foundation of SCD care centered around the communities rather than just the mother. This foundation of care works through existing social-cultural structures and does not conflict with the local gendered norms and values within families. The second level will be identifying modes of integration of these networks of care in SCD care provision. This is crucial to ensure that those responsible for care are equipped with the necessary knowledge and skills to manage SCD at different levels. One way of ensuring the family members is knowledgeable with SCD is to provide regular health education sessions during the SCD clinics. The survey done in this study has shown that 12% of the children are brought to the clinics by extended family members. A study done on the same settings amongst pregnant women attending antenatal clinics has shown that only 14.7% out of 600 women who participated had good knowledge on SCD (Tutuba et al., 2022). This shows the gap in knowledge on the general understanding of SCD even amongst communities with the highest burden of the disease. Developing structured health education programs that can be offered using local languages complemented with pictorial presentations targeting these large networks of care may help steer interests to understand and learn more about SCD (Bukini, Mbekenga, Nkya, Purvis, et al., 2020). Developing counselling programs to capture other family members will be a huge advantage for communities who already have high prevalence of SCD (Anie et al., 2016). The larger clinical goal of Newborn Screening (NBS) programs for SCD is to improve quality of life of children born with SCD. However, this can only be achieved if we properly consider ways to include the social experiences of the patients and their caregivers in the clinical care (Bukini et al., 2020; Green et al., 2016). Locally relevant ethical theories, such as “Ubuntu”, may help us to understand ways to formulate the design of SCD programs that are rooted in the patient lived experiences (Behrens, 2013; Chuwa, 2012; Tumani Chuwa, 2014) Also redefining the limitations of care practices that may not be well applicable in sub-Saharan settings will help to identify or address the unique structural and ethical conundrums that are unique in these settings. Through focusing on health education and counselling programs that only recognizes parents as the primary caregivers we are leaving out a great support system that already exist as part of the childcare. Diagnosis of SCD in one member of a family may indicate that there ‘*may be’* other family members either with the SCD or with SCD trait. It is undoubtedly paramount for countries in Sub-Saharan Africa with the highest burden of SCD to invest in health education and counselling programs as the most basic approach to facilitate communication and knowledge sharing between families and clinicians. But of more importance is to also think of how the programs will have high impact to the communities affected with the disease. SCD programs that need to be developed in the contexts of Africa must be able to identify and address the structural as well as understanding the social contexts of the communities (Anie et al., 2016; Dennis-Antwi et al., 2011; Schmidtke & Cornel, 2019).

## Conclusion

The analysis done in this study have shown how the existing networks of care within our communities have been crucial in supporting care of children with SCD in the settings. Research is needed to further provide recommendations on the inclusion of the informal support in health care provision. Other future work that needs to be explored is recognizing the practical value of Ubuntu principle and more specifically its translation into clinical care. While SCD care has been used to substantiate the importance of the networks of care, we argue that the proposed approach can be applicable to other public health programs designed for chronic illnesses.

## Data Availability

All data produced in the present work are contained in the manuscript

## Ethical Consideration

The study was approved by the Muhimbili University of Health and Allied Sciences Research Ethics Committee (Ref. No. 2017-10-20/ AEC/ Vol. XII/85). Consent was sought prior to the conduct of the study

## Acknowledgements

We thank parents and caregivers who have participated in this research, and the Sickle Cell Programme at Muhimbili University of Health and Allied Sciences. We also thank staff and administration of Regional Referral Hospitals in Dar es Salaam where data was collected in the SCD clinics. Special thanks to Ruth Lwakatare for designing the infographic to illustrate the survey results.

## Funding

Research funding for this work was supported through student small grant at MUHAS

## Competing Interest

Authors declare no conflict of interests

## Authors contribution

DB designed the study, interacted with participants, performed interviews, assisted in transcriptions and translations, analysis and drafting of the manuscript. KM, JM and CM supervised DB and reviewed the drafts of the manuscripts. IW, CK, JO, DM assisted with data collection and reviewing the manuscript. RL, UM assisted in analyzing the survey data and revising the manuscript. All authors reviewed and approved the final manuscript.

## Data Availability

All data relevant to the study are included in the article.

## Notes

### Competing Interest Statement

The authors have declared no competing interest.

### Funding Statement

The study was funded by the Sickle Cell Program at Muhimbili University of Health and Allied Sciences

### Author Declarations

Ethics Committee of Muhimbili University of Health and Allied Sciences gave ethical approval for this work

